# Loneliness is associated with 15-year risk of stroke: The Framingham Heart Study

**DOI:** 10.64898/2026.11.12.26367982

**Authors:** Oluchi Ekenze, Ayantika Banerjee, Vasileios-Arsenios Lioutas, Dibya Himali, Sudha Seshadri, Jose R. Romero, Hugo J. Aparicio, Alexa Beiser, Joel Salinas

**Affiliations:** NHLBI’s Framingham Heart Study, Framingham, MA; Department of Neurology, Boston University Chobanian and Avedisian School of Medicine, Boston, MA; Department of Neurology, Beth Israel Deaconess Medical Center, Harvard Medical School, MA; The Glenn Biggs Institute for Alzheimer’s and Neurodegenerative Diseases, University of Texas Health Sciences Center, San Antonio, TX; Department of Biostatistics, Boston University School of Public Health, Boston, MA; Department of Neurology, New York University Grossman School of Medicine, NY

## Abstract

Psychosocial factors are promising candidates as modifiable risk factors for stroke. Loneliness is of interest given its rising prevalence worldwide; yet the temporal relationship of loneliness with subsequent stroke remains unclear.

**Aim:** To clarify the association between timing of loneliness and incident stroke.

**Methods:** Framingham Heart Study participants free of stroke with at least two assessments of loneliness prior to incident stroke (at baseline exam, and at prior exam up to 15 years before baseline) were included. Loneliness was assessed using response to the “I felt lonely” item on the Center for Epidemiologic Studies Depression Scale, defined as feeling lonely for ≥3 days within the past week. The recency of loneliness was defined as prior loneliness (present at the prior exam) and baseline loneliness (present at the baseline exam). The primary analysis tested separately for associations between prior loneliness vs no prior loneliness, baseline versus no baseline loneliness, and incident stroke, with 15 years of follow up. Cox models were adjusted for age and sex (model 1) and additionally vascular risk factors (model 2), depressive symptoms (model 3), and physical activity (model 4). Interactions between loneliness and age and sex were explored.

**Result:** Among 4307 participants (mean age 62±10 years, 55% women), 198 (4.5%) had an incident stroke over a median 6.5-year follow-up period. Prior loneliness was reported by 177 (4.1%) and 226 (5.2%) reported baseline loneliness. Compared to participants without prior loneliness, those with prior loneliness had nearly 3-fold increase in stroke risk after adjustment for age, sex, vascular risk factors, depressive symptoms, and physical activity (HR=2.85, 95% [CI] 1.51-5.39). The effect was primarily observed in participants <80 years (mean age 60 years.HR=3.21, 95% CI 1.69-6.13). No increased risk of stroke was observed among participants with loneliness at baseline.

**Conclusion:** Our findings support consideration of loneliness as a potentially modifiable risk factor, particularly in midlife, for incident stroke.

**3-5 Bullet points:**

- Loneliness, reported on average 7.6 years prior to a baseline exam, was associated with long-term risk of incident stroke
- The increased risk of stroke associated with prior loneliness was observed independent of vascular risk factors, the presence of depressive symptoms, or physical activity status
- The association between prior loneliness and incident stroke was observed primarily in individuals <80 years, around a mean age of 60 years, representing an independent risk factor that may be present at mid-life
- Efforts to address loneliness, early in the life course, may reduce future stroke incidence

## Introduction

Social connections integrate the structural and functional aspects of interpersonal relations.^1^ Loneliness is a negative subjective evaluation of the quality of those connections and the support received from them.^2^ For example, an individual can experience loneliness based on whether they feel supported, or by their satisfaction with the quality of their social contacts. Loneliness is common; approximately one-third of the adult US population >45 years old and approximately 43% of adults >60 years old report loneliness.^1,3^ Loneliness is associated with a range of negative health outcomes, including dementia,^4,5^ sleep disturbances,^6^ coronary heart disease,^7^ stroke,^8–10^ and cardiovascular disease.^11^ Further, loneliness has been linked to detrimental behavioral changes, such as physical inactivity^12,13^ and smoking,^13,14^ and with depression.^15^ Loneliness may influence biological pathways that lead to atherosclerosis,^16,17^ hypertension,^16,18^ and metabolic disease.^19–21^

Few population-based studies have evaluated the prospective relationship between loneliness and incident stroke.^8,9^ Using data from the UK Biobank,^9^ investigators observed that over a 7-year follow up period lonely participants had a higher risk of incident stroke after controlling for age, sex, diastolic and systolic blood pressure, smoking, physical inactivity, and alcohol consumption. More recently, investigators in the Health and Retirement Study,^8^ over a 10-12 year follow up period, observed that loneliness was associated with stroke in older participants. Importantly, the impact of loneliness may differ based on its timing and chronicity.^10,22,23^ Data is lacking, however, to understand the temporal association between loneliness and stroke. The Health and Retirement study observed that persistent loneliness over time (6–8-year interval period) was associated with incident stroke despite controlling for vascular and lifestyle risk factors^8^. Because loneliness is a potentially modifiable psychosocial risk factor, there is a need for longitudinal studies among community-based individuals to better understand the associations of loneliness, and changes in loneliness status over time, with incident stroke.

Our objective was to address this knowledge gap using comprehensive loneliness and stroke incidence data from the prospective, observational Framingham Heart Study (FHS). We assessed whether the timing and persistence of loneliness predict incident stroke over a fifteen-year follow-up period. We hypothesized that individuals with longer duration of loneliness would have higher risk of incident stroke and that the association would be independent of conventional stroke risk factors.

## Methods

### Sample

The recruitment of participants in the FHS was previously described.^24^ Briefly, the FHS is a multi-generational cohort begun in 1948 with the recruitment of the Original Cohort. In 1971, the offspring of the Original Cohort (Offspring Cohort) and their spouses were recruited. The Third-Generation Cohort (Gen 3 Cohort), including grandchildren of the Original Cohort, were recruited in 2002 along with the NOS cohort (spouses of members of the Offspring Cohort who were not in the original Offspring Cohort). To reflect the increased racial and ethnic diversity of the town of Framingham, Massachusetts, recruitment for the Omni 1 cohort was initiated in 1994 and later the Omni 2 cohort along with the Gen 3 and NOS cohorts. FHS participants were examined on average every 2-8 years (Original every 2 years; Offspring and Omni 1 every 4-8 years; Gen 3, NOS, and Omni 2 every 6 years), including medical history, physical examination, anthropometry, assessment of psychosocial factors by questionnaire, and 12-lead electrocardiography.

In the current study, we included all FHS participants with at least two assessments for loneliness using the response to the loneliness item in the Center for Epidemiologic Studies Depression Scale (CES-D) before the occurrence of incident stroke. Among 5987 participants who attended baseline examinations, 5899 had data on loneliness. Participants were excluded if they did not have at least two assessments for loneliness, were aged less than 45 years, had prevalent stroke at baseline, did not have follow up assessment to determine the occurrence of stroke, or had missing covariates (Figure 1). The final sample consisted of 4307 individuals, including 3936 aged less than 80 years and 371 aged at least 80 years.

**Figure 1:**
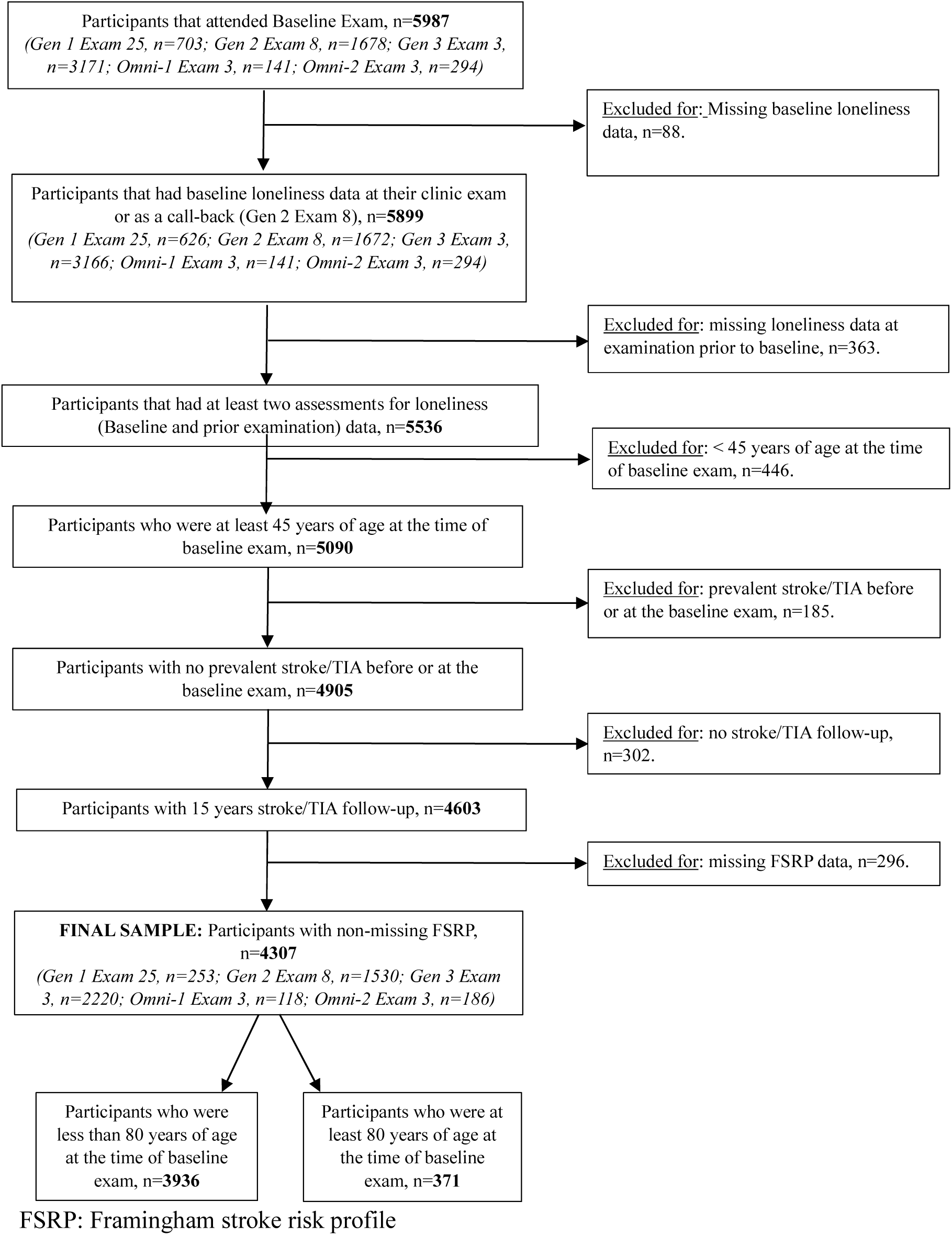
Sample flow chart of study participants.

### Exposure

Participants completed the self-reported CES-D, which is a questionnaire assessing frequency of each of a series of twenty depressive symptoms in the past week. Symptoms are reported as occurring “rarely or none of the time (less than 1 day)” (0 points), “some or a little of the time (1-2 days)” (1 point), “occasionally or a moderate amount of time (3-4 days)” (2 points), or “most or all of the time (5-7 days)” (3 points). Consistent with prior FHS studies^5,25,26^, loneliness was assessed using the response to the question “I felt lonely” and based on a scale of number of days feeling lonely within the past week. We defined loneliness as feeling lonely for ≥ 3 days within the past week. We assessed loneliness at two-time points: prior to baseline and at baseline (median interval 7.6 years [interquartile range 7.0 – 8.0 years]). We assessed baseline loneliness at exam 25 for the Original Cohort (1997-1999), exam 8 for the Offspring Cohort (2005-2008), and exam 3 for the Omni 1 (2007-2008), Gen 3 (2016-2019), and Omni 2 (2016-2019) cohorts. Prior loneliness was assessed at exam 23 for the Original Cohort (1992-1996), exam 7 for the Offspring cohort (1998-2001), and exam 2 for the Omni 1 (1999-2001), Gen 3 (2008-2011), and Omni 2 (2009-2011) cohorts.

Loneliness patterns over time have been described in four mutually exclusive groups^5^: (a) no loneliness (participants did not report loneliness at either the prior exam or the baseline exam); (b) incident loneliness (participants report loneliness only at the baseline examination); (c) transient loneliness (participants who reported loneliness only at the exam prior to baseline); and, (d) persistent loneliness (participants who reported loneliness at both the prior and baseline examinations). The sample sizes of these four loneliness trajectories are presented in Supplementary Table 1A. Our central question regarding the impact of loneliness was focused on whether the recency of loneliness is associated with incident stroke. Recency of loneliness may be relevant because studies show that recent psychosocial stress is associated with increased risk of stroke.^27^ We grouped these participants into two time-window exposures (Figure 2):

- **Prior loneliness:** loneliness reported distal to the examination immediately preceding baseline (including transient and persistent cases)
- **Baseline loneliness:** loneliness reported proximal to the baseline examination (includes incident and persistent cases)

**Figure 2.**
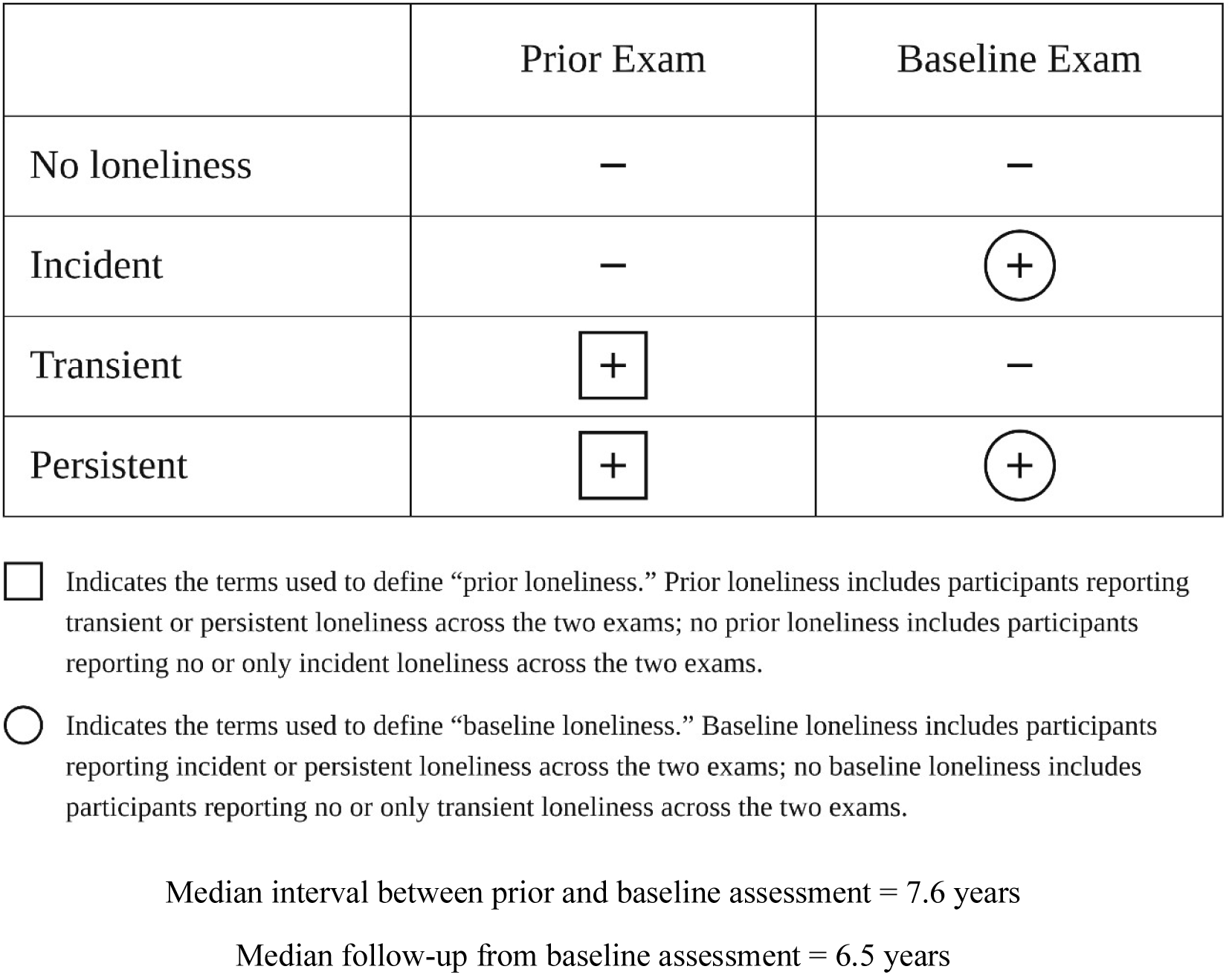
Summary of terms used to describe and analyze prior and baseline loneliness status over time.

All analyses used these intentionally overlapping windows as the primary exposure.

### Outcome

Ongoing monitoring and observation for incident stroke cases includes day-to-day screening of participants admitted to the local hospital in Framingham or other area hospitals, biennial (Original) or quadrennial (other cohorts) FHS examinations, yearly health updates, continuing appraisal of Medicare entitlements data, or description by participants, their family members, or primary care physicians through a stroke pager (1-800-FHSTROK). All health information for any emergency department visits, admissions, or death is evaluated to complement thorough screening.

Adjudication of stroke and transient ischemic attack (TIA) is performed by a panel of at least two FHS vascular neurologists who review all records, including medical information, imaging studies (head CT/MRI, CTA/MRA, conventional angiography), and diagnostic testing done to assess stroke etiology. A stroke event is defined as a sudden or rapid onset of a focal neurological deficit of vascular origin that persists for greater than 24 hours. Stroke cases include ischemic stroke, intracerebral hemorrhage, and subarachnoid hemorrhage. TIA is defined as a similar event persisting for less than 24 hours, with complete resolution of symptoms. Information on acute stroke treatment received, assessment for cardio-embolism (electrocardiogram, transthoracic or transesophageal echocardiography, cardiac event monitor, implantable loop recorder), carotid ultrasound, and autopsy (where applicable) are reviewed. Neuroimaging studies (head CT/MRI) are available in more than 90% of cases. In the present study, we included all stroke and TIA cases.

The follow up period extended from baseline exam up to 15 years (median 6.5 years).

Censoring for participants who did not experience an incident stroke occurred at the last date known to be stroke free, end of study period (15-year follow up), or death from other causes.

The study was approved by the Institutional Review board of Boston University Medical Center.

### Covariates

The covariates in the study were obtained at baseline exam and include the revised Framingham Stroke Risk Profile (FSRP), a research and clinical tool that incorporates an individual’s clinical characteristics and risk factors to estimate 10-year risk of stroke.^28^ The FSRP incorporates the effects of the following risk factors: age, sex, systolic blood pressure, use of antihypertensive medication, prevalent cardiovascular disease (coronary artery disease, congestive heart failure, or peripheral vascular disease), diabetes mellitus, cigarette smoking, and prevalent atrial fibrillation. Similar to prior FHS studies of loneliness using the CES-D measure,^4,5^ depressive symptoms were assessed using CES-D scores modified to exclude the loneliness item to derive a depression score independent of loneliness. The modified CES-D score was used as a continuous variable and was natural log transformed to normalize its distribution. Physical activity index was assessed using information from a structured questionnaire and defined as a composite score based on the number of hours spent per day on an activity and weight-based oxygen consumption required for that activity.^29^ The Physical Activity Index was available for the Offspring, third-generation, Omni 1 and Omni 2 cohorts.

### Statistical Analysis

Baseline characteristics were obtained for the overall sample, in those with or without prior loneliness and with/without baseline loneliness. We also stratified our sample based on age (< and ≥ 80 years). We chose this age cutoff a priori because previous research in the FHS and others has shown difference in the burden of loneliness in the two age strata.^4,30^

We summarized continuous variables as mean (SD) or median [Q1, Q3], and categorical variables as number (%). The primary analysis examined the association between loneliness and risk of incident stroke, separately by the recency of loneliness (prior loneliness vs no prior loneliness; baseline loneliness vs no baseline loneliness). Secondary analysis related recency of loneliness to incident stroke with a less conservative threshold for loneliness status, defined as a report of feeling lonely for at least 1 day in the past week.

Four multivariable Cox proportional hazard regression models were evaluated to obtain hazard ratios (HR) and 95% confidence intervals (CI). The primary model adjusted for age and sex. The second model was additionally adjusted for FSRP score. A third model additionally adjusted for depressive symptoms using the modified CES-D score excluding the loneliness item. Finally, a fourth model additionally adjusted for physical activity index.

We assessed the interaction of loneliness with age and sex. A threshold of p value <0.1 was used to identify significant interaction and we performed stratified analyses as indicated. For all other analysis, a two-sided p-value < 0.05 was considered statistically significant. Statistical analyses were performed using SAS version 9.4 (SAS Institute Inc.).

## Results

Demographic and clinical characteristics of the study participants (overall, stratified by loneliness status, and those excluded) are shown in Table 1. The study included 4307 participants (median age 61years, inter quartile range [IQR] 55, 68). Overall, 177 of 4307 (4.1%; median age 61 years [IQR 65, 69]) had prior loneliness while 226 (5.2%; median age 62 years [IQR 56,71]) had loneliness at baseline. The excluded participants (n=1680, median age 57 years [IQR: 44, 80]), had more prevalent cardiovascular disease than the overall included sample, but similar median FSRP score and proportion of participants with hypertension. Supplementary tables 1A &1B shows the number of participants with loneliness and incident stroke in each age group.

**Table 1:** Baseline characteristics of study participants.

|  |  | **Prior loneliness ≥3 days/week** |  | **Baseline loneliness ≥3 days/week** |  | **Excluded participants** |
|---|---|---|---|---|---|---|
|  | **Total**<br>**N=4307** | **No prior loneliness**<br>**N=4130** | **Prior loneliness**<br>**N=177** | **No baseline loneliness**<br>**N=4081** | **Baseline loneliness**<br>**N=226** | **N=1680** |
| Women, n (%) | 2369 (55.00) | 2253 (54.55) | 116 (65.54) | 2237 (54.81) | 132 (58.41) | 976 (58.10) |
| Age, years, median [Q1, Q3] | 61 [55, 68] | 61 [55, 68] | 61 [56, 69] | 61 [55, 68] | 62 [56, 71] | 57 [44, 80] |
| ***FHS Cohort, n (%)*** |  |  |  |  |  |  |
| Original | 253 (5.87) | 232 (5.62) | 21 (11.86) | 235 (5.76) | 18 (7.96) | 450 (26.79) |
| Offspring | 1530 (35.52) | 1488 (36.03) | 42 (23.73) | 1436 (35.19) | 94 (41.59) | 148 (8.81) |
| Gen 3 | 2220 (51.54) | 2125 (51.45) | 95 (53.57) | 2125 (52.07) | 95 (42.04) | 951 (56.61) |
| Omni 1 | 118 (2.74) | 110 (2.66) | 8 (4.52) | 113 (2.77) | 5 (2.21) | 23 (1.37) |
| Omni 2 | 186 (4.32) | 175 (4.24) | 11 (6.21) | 172 (4.21) | 14 (6.19) | 108 (6.43) |
| ***Vascular Risk Factors*** |  |  |  |  |  |  |
| Systolic blood pressure, mean (SD), mmHg | 125 (16) | 125 (16) | 123 (17) | 125 (16) | 125 (16) | 125 (19) |
| Diastolic blood pressure, mean (SD), mmHg | 75 (10) | 75 (10) | 75 (9) | 75 (9) | 74 (10) | 73 (10) |
| Total cholesterol, mean (SD), mg/dL | 189.27 (36.60) | 189.11 (36.59) | 193.16 (36.76) | 189.20 (36.41) | 190.61 (40.07) | 186.17 (36.86) |
| Total cholesterol/HDL ratio, median [Q1, Q3] | 3.30 [2.69,4.10] | 3.31 [2.70,4.09] | 3.29 [2.59,4.30] | 3.31 [2.69,4.10] | 3.190 [2.69,4.16] | 3.35 [2.70, 4.27] |
| Hypertension, n (%) | 1944 (45) | 1865 (45) | 79 (45) | 1837 (45) | 107 (47) | 689 (41) |
| Treatment for hypertension, n (%) | 1561 (36) | 1497 (36) | 64 (36) | 1468 (38) | 93 (41) | 541 (33) |
| Current smoker, n (%) | 270 (6) | 249 (6) | 21 (12) | 247 (6) | 23 (10) | 133 (8) |
| Atrial fibrillation, n (%) | 194 (4) | 187 (5) | 7 (4) | 179 (4) | 15 (7) | 125 (7) |
| Diabetes mellitus, n (%) | 470 (11) | 446 (11) | 24 (14) | 436 (11) | 34 (15) | 100 (8) |
| Diabetes treatment, n (%) | 349 (8) | 331 (8) | 18 (10) | 321 (8) | 28 (13) | 100 (6) |
| Lipid lowering medication use, n (%) | 1393 (32) | 1345 (33) | 48 (27) | 1328 (33) | 65 (29) | 300 (18) |
| Prevalent cardiovascular disease, n (%) | 354 (8) | 330 (8) | 24 (14) | 327 (8) | 27 (12) | 362 (22) |
| Modified CESD score (range 0–57)<br>(continuous, excluding loneliness item),<br>median [Q1, Q3] | 4.00 [2.00,9.00] | 4.00 [2.00,8.00] | 10.00 [6.00,19.00] | 4.00 [1.11,8.00] | 4.00 [2.00,9.00] | 5.00 [2.00, 10.00] |
| FSRP, median [Q1, Q3] | 0.02 [0.01,0.06] | 0.02 [0.01,0.06] | 0.02 [0.01,0.07] | 0.02 [0.01,0.06] | 0.02 [0.01,0.08] | 0.01 [0.00,0.02] |
| Physical activity index, median [Q1, Q3] | 33.40 [30.70,36.70] | 33.50 [30.70,36.70] | 32.40 [29.80.70,35.60] | 33.40 [30.70,36.70] | 32.90 [29.60,36.60] | 33.00 [30.20,36.20] |

Values are mean (SD) for continuous variables that are normally distributed and median [1<sup>st</sup> quartile, 3<sup>rd</sup> quartile] for continuos variables that have a skewed distribution. Values are number (%) for categorical variables.

The median follow-up was 6.5 years [IQR: 4.2, 13.3] in all participants, 6.3 years [IQR: 4.1, 13.6] in those <80 years and 8.3 years [IQR: 5.5, 12.0] in those ≥80 years) (Supplementary Table 2). Among all participants, 14/177 (7.9%) with prior loneliness, and 11/226 (4.9%) with loneliness at baseline had an incident stroke. Among those aged <80 years, 12/154 (7.8%) with prior loneliness and 10/195 (5.1%) with baseline loneliness subsequently had incident stroke. In participants ≥80 years, only 2/23 (8.7%) with prior loneliness and 1/31 (3.2%) with loneliness at baseline had an incident stroke (Supplementary Table 1B).

We observed significant interactions between age (<80 versus ≥80) and each of prior and baseline loneliness (p = 0.02 and p=0.07, respectively) in their associations with incident stroke. There was no interaction between sex and either prior loneliness (p=0.6) or baseline loneliness (p=0.17).

### Relation between recency of loneliness and incident stroke

#### All participants

Compared to those without prior loneliness, participants with loneliness at the prior examination had a 101% higher risk of incident stroke (hazard ratio [HR] 2.01; 95% CI 1.16, 3.48) in models adjusted for age and sex (Table 2). The results persisted after additionally adjusting for FSRP score in model 2 (HR 1.93; 95% CI 1.11, 3.34), depressive symptoms (modified CES-D) in model 3 (HR 1.84; 95% CI 1.05, 3.23), and physical activity index in model 4 (HR 2.85; 95% CI 1.51, 5.39). Conversely, loneliness at baseline was not associated with the risk of incident stroke. Cumulative incidence curves of stroke in participants with or without prior loneliness are shown in Figure 3.

**Figure 3:**
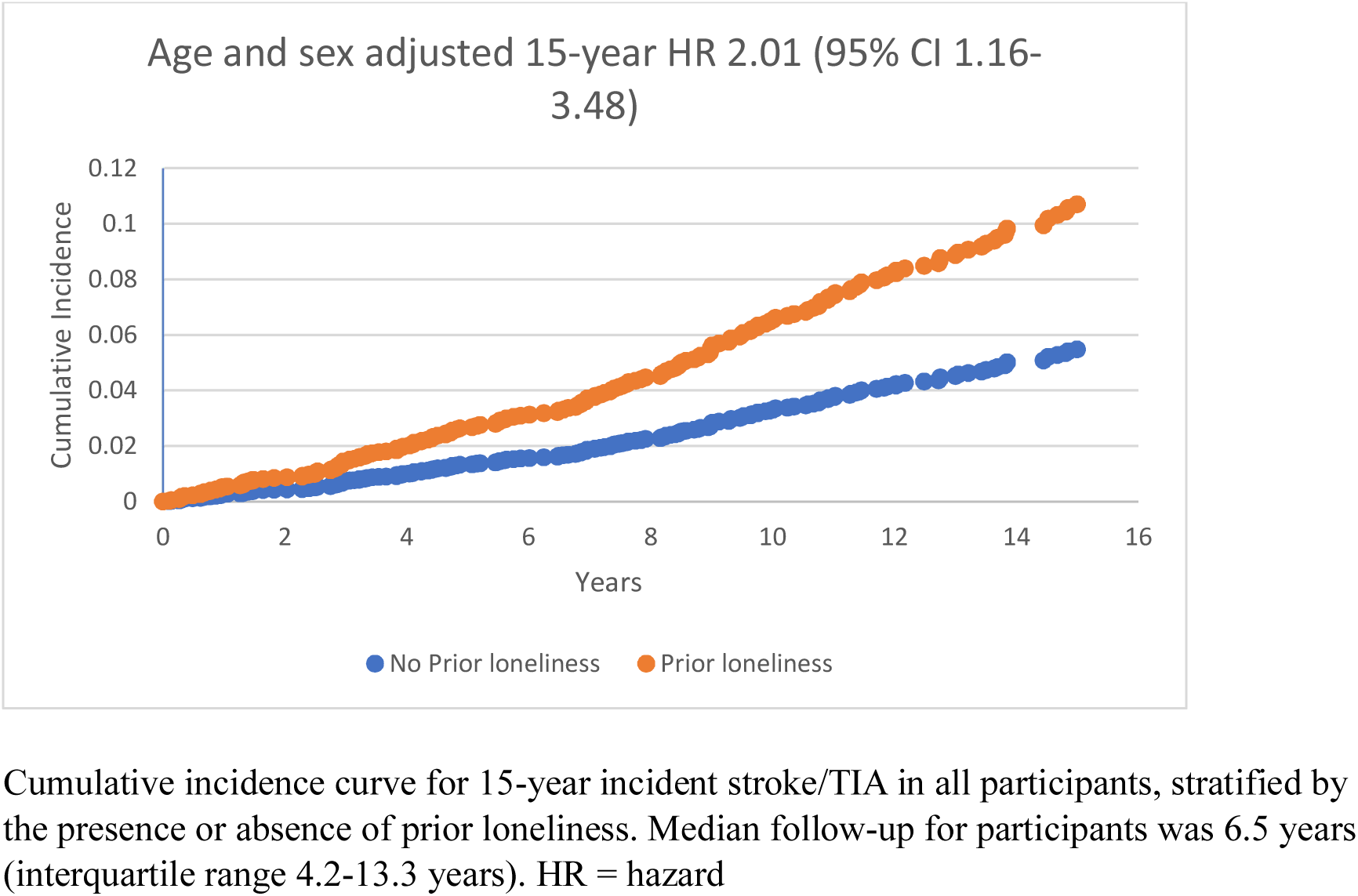
Fifteen-year cumulative incidence of stroke/TIA, stratified by self-report of prior loneliness defined as loneliness ≥3 days/week.

**Table 2:** Cox-proportional hazard regression of incident stroke/TIA by loneliness status ≥3 days/week (all participants).

|  | **Model 1 (n=4307)** |  |  | **Model 2 (n=4307)** |  |  | **Model 3 (n=4307)** |  |  | **Model 4 (n=4009)<sup>#</sup>** |  |  |
|---|---|---|---|---|---|---|---|---|---|---|---|---|
|  | ***Stroke/TIA cases/n*** | ***HR (95% CI)*** | ***P-value*** | ***Stroke/TIA cases/n*** | ***HR (95% CI)*** | ***P-value*** | ***Stroke/TIA cases/n*** | ***HR (95% CI)*** | ***P-value*** | ***Stroke/TIA cases/n*** | ***HR (95% CI)*** | ***P-value*** |
| Prior Loneliness |  |  |  |  |  |  |  |  |  |  |  |  |
| No | 184/4130 | 1.00 (ref) |  | 184/4130 | 1.00 (ref) |  | 184/4130 | 1.00 (ref) |  | 142/3854 | 1.00 (ref) |  |
| Yes | **14/177** | **2.01 (1.16,3.48)** | **0.013** | **14/177** | **1.93 (1.11,3.34)** | **0.019** | **14/177** | **1.84 (1.05,3.23)** | **0.033** | **11/155** | **2.85 (1.51,5.39)** | **0.001** |
| Baseline Loneliness |  |  |  |  |  |  |  |  |  |  |  |  |
| No | 187/4081 | 1.00 (ref) |  | 187/4081 | 1.00 (ref) |  | 187/4081 | 1.00 (ref) |  | 142/3804 | 1.00 (ref) |  |
| Yes | 11/226 | 0.88 (0.48,1.62) | 0.680 | 11/226 | 0.88 (0.48,1.61) | 0.673 | 11/226 | 0.78 (0.42,1.47) | 0.444 | 11/205 | 1.09 (0.57,2.08) | 0.800 |

Model 1: adjusted for age and sex
Model 2: adjusted for age, sex, Framingham Stroke Risk Profile score (FSRP)
Model 3: adjusted for age, sex, FSRP, modified Center for Epidemiologic Studies Depression Scale (continuous, logarithm of CES-D excluding loneliness item)
Model 4: adjusted for adjusted for age, sex, FSRP, modified CES-D, physical activity index
#model 4 done in Offspring, Third Generation, Omni 1 and 2 cohorts

#### Participants <80 years

Among those <80 years, and in models adjusted for age and sex, participants with prior loneliness had a 252% higher risk of incident stroke (HR 3.52; 95% CI 1.94, 6.40) compared to those without loneliness at the prior examination (Table 3). The results persisted after additionally adjusting for FSRP score (HR 3.47; 95% CI 1.91, 6.30), depressive symptoms (HR 3.17; 95% CI 1.70, 5.89), and physical activity index (HR 3.21; 95% CI 1.69, 6.13). Conversely, loneliness at baseline was not associated with the risk of incident stroke in this age group. Survival curves for cumulative incidence of stroke by prior loneliness in those aged <80 years are shown in Supplementary Figure 1.

**Table 3:**
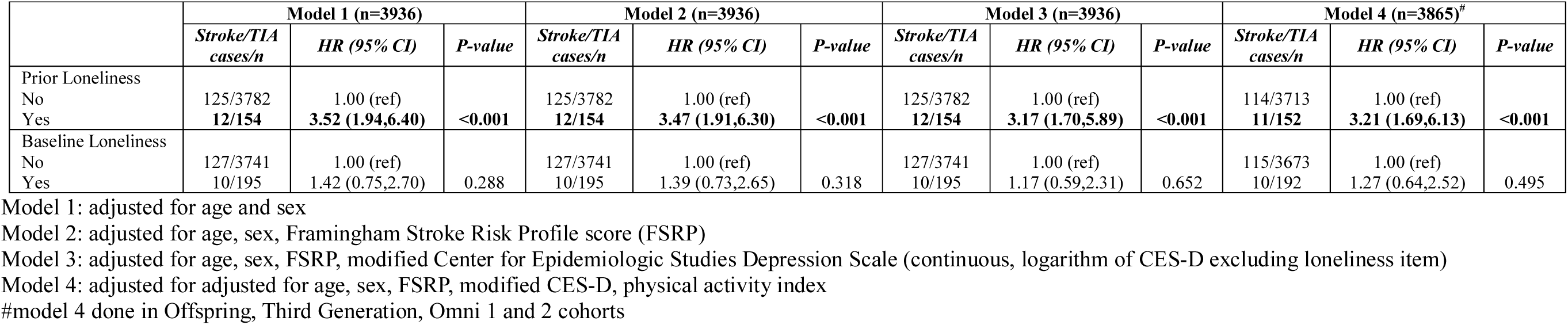
Cox-proportional hazard regression of incident stroke/TIA by loneliness status ≥3 days/week (age <80 years).

#### Participants ≥80 years

We did not observe associations between prior loneliness (HR 0.60; 95% CI 0.14, 2.47) or baseline loneliness (HR 0.21; 95% CI 0.03, 1.51) with incident stroke in models adjusted for age and sex (Supplementary Table 3). Due to the small number of participants in this subgroup of older adults and small number of strokes among them, we did not perform further analyses in additional models.

#### Secondary analysis

In secondary analysis using a less conservative definition for loneliness as feeling lonely ≥1 day in the past week, across all age strata, in models adjusted for age and sex, participants with prior loneliness had a 51% higher risk of incident stroke (HR 1.51; 95% CI 1.06, 2.15) compared to those without prior loneliness (Supplementary Table 4). The results persisted after additionally adjusting for FSRP score (HR 1.51; 1.06, 2.15), depressive symptoms (HR 1.84; 95% CI 1.05, 3.23) and physical activity index (HR 1.60; 95% CI 1.06, 2.41). Conversely, loneliness at baseline was not associated with the risk of incident stroke.

Among those <80 years, compared to those who did not have prior loneliness (≥1 day/week), participants with prior loneliness had a 76% higher risk of incident stroke (HR 1.76; 95% CI 1.17, 2.67) in models adjusted for age and sex (Supplementary Table 5). The results persisted after additionally adjusting for FSRP score (HR 1.76; 1.17, 2.66), depressive symptoms (HR 3.17; 95% CI 1.70, 5.89) and physical activity index (HR 1.69; 95% CI 1.08, 2.64). Loneliness at baseline was not associated with the risk of incident stroke in this age group.

In those aged ≥80 years, we did not observe an association between prior loneliness (≥1 day/week) and incident stroke, after adjusting for age and sex (HR 1.14; 95% CI 0.57, 2.27). No significant associations were observed in subsequent models. Loneliness at baseline was not associated with incident stroke in those aged ≥80 years (Supplementary Table 6). Using loneliness definition (≥1 day/week) and with larger number of participants and stroke events in this subgroup of older participants, similar results were obtained with no effect in those ≥ 80 years.

## Discussion

In this community-based sample, participants with prior loneliness (median 7.6 years prior to baseline), defined as feeling lonely at least three days in the past week, had an increased risk of stroke over a follow up of 15 years in the overall sample and among participants less than 80 years of age. The increased risk was present after additional adjustment for baseline demographics, vascular risk factors, self-reported symptoms of depression, and physical activity.

Our findings on the association between loneliness and incident stroke are aligned with earlier studies that noted an increased risk of stroke in lonely individuals.^8,31^ Our study expands on this knowledge, suggesting a dose-response relationship; in fully adjusted models, we observed a 3-times higher risk of stroke in those experiencing loneliness ≥3 days in the past week (185% increase in stroke risk) compared to ≥1 day in the past week (60% increase in stroke risk). In addition, while prior studies showed that chronic loneliness was associated with a 25% higher risk of stroke,^8^ the current study observed, a stronger effect with a 185% higher risk overall, and a 221% higher risk in those aged <80 years. Differences in the increased risk may be due to different populations that were studied. The current study was in a younger population (mean age 62 years) while a previous study^8^ involved an older group with a mean age of 67 years.

Our results suggest that the impact of loneliness on risk of incident stroke may be more pronounced in younger age groups; we observed no association between loneliness and incident stroke in those aged ≥80 years, Further study of this association (beyond age and sex adjusted analyses) among older individuals was limited by the small number of strokes in this age group. However, we note that using the definition of loneliness as ≥1 day/week, the overall number of stroke events increased in those ≥80 years, but the same pattern was observed with no effect of baseline loneliness or prior loneliness with risk of stroke in those ≥ 80 years. According to a random survey of Medicare-insured adults,^32^ older adults ≥80 years reported more loneliness than younger adults. However, older adults have a greater number of prevalent vascular risk factors,^33^ so loneliness may play a relatively larger contributory role among younger individuals in relation to stroke risk. Loneliness is also associated with dementia in people at least 60 years of age,^4,34^ suggesting that the impact of loneliness on brain health and in association with increased risk of neurologic disease may begin as early as midlife.^35,36^

Our results reinforce previous findings that the timing of loneliness, and not merely the presence or absence of loneliness, may influence risk of subsequent stroke. Loneliness may induce stress,^17,37^ leading to activation of the hypothalamic pituitary axis and sympathetic adrenomedullary axis,^17,19^ resulting in a rise in blood pressure,^17,18^ endothelial dysfunction,^19,37,38^ decreased glucocorticoid receptor sensitivity^19^ and accelerated atherosclerosis^17,38,39^ leading to increased risk of stroke. Loneliness also leads to exaggerated autonomic response with increased total peripheral arterial resistance and reduced heart rate variability, which contributes to hypertension, ischemic heart disease, and poor cardiac output.^40^ Further, stressful conditions like chronic loneliness are associated with release of cortisone, and a rise in blood glucose.^41^ In addition, loneliness has been associated with release of inflammatory mediators such as interleukin-6, interleukin-1β, tumor necrosis factor-alpha and natural killer cells causing a state of chronic inflammation, endothelial dysfunction and contributing to atherosclerosis,^40^ likely increasing the risk for cardiovascular diseases^42^ including stroke.^43,44^ Loneliness may lead to atherogenic behaviors such as tobacco use,^13,14^ alcohol abuse, low diet quality including low fruit and vegetable intake,^45^ maladaptive eating behavior^46^ and physical inactivity.^12^ Further, previous population-based studies have identified an association between loneliness and depression^47^ a risk factor for stroke.^48^ Our analyses observed that loneliness remained an independent risk factor for stroke after adjustment for depressive symptoms.

The net effect of the physiological, behavioral, metabolic, inflammatory, and psychological changes from loneliness may lead to increased risk for stroke. These relationships with loneliness are likely to occur gradually over years^18^ in a dose-response fashion^49^ and, particularly relevant to the present study, may explain observed associations of prior loneliness but not loneliness at baseline with incident stroke, and higher risk of stroke among those reporting loneliness ≥3 days in the past week compared to ≥1 day. The association of loneliness with incident stroke persisted after adjusting for vascular risk factors, depressive symptoms, and physical inactivity, suggesting that these vascular, psychiatric, and behavioral pathways only partially explain the link between loneliness and stroke.

The strengths of the present study lie in the large sample size of the primary analytic sample, robust longitudinal clinical phenotyping, and the ability to assess loneliness at two different points over time. There were also limitations. The population of our cohort was predominantly White adults, reducing the generalizability of our results to a more diverse population. The smaller sample size among older adults restricted further analysis in this age group over age 80 years. Also, the use of the single-item self-reported response to the lonely question may have led to an underestimation of loneliness, however assessing loneliness using the single item in the CES-D related to loneliness has showed substantial correlation with the revised UCLA loneliness scale.^50^ Participants may under-report loneliness because of social stigma; however, previous work has shown consistent concordance using this and other loneliness assessment tools.^50^

## Conclusion

Our findings support the role of loneliness as a temporally-sensitive risk factor for incident stroke in community-dwelling individuals, primarily in those younger than age 80. Further investigating loneliness as a possible early protective target to reduce incident stroke risk is merited.

## Public health Implications

Loneliness is potentially modifiable, is feasible to assess in clinical practice, and may have an important role in stroke prevention particulary in midlife by identifying individuals at increased risk of stroke. Measures aimed at reducing loneliness may help decrease the burden of stroke.

## Author Contributions

Design and conceptulaization of the study: OE, JRR, AB, HJA, JS

Major role in the acquisition of data: AB, HJA, JS

Analysis or interpretation of data: OE, AB, AB, HJA, JS

Drafting or revisisng the manuscript for intellectual content: OE, AB, VL, DH, SS, JRR, AB, HJA, JS.

## Data Availability

Requests to access de-identified data from the Framingham Heart Study can be made at https://www.framinghamheartstudy.org/fhs-for-researchers

---

**Supplementary Figure 1:**
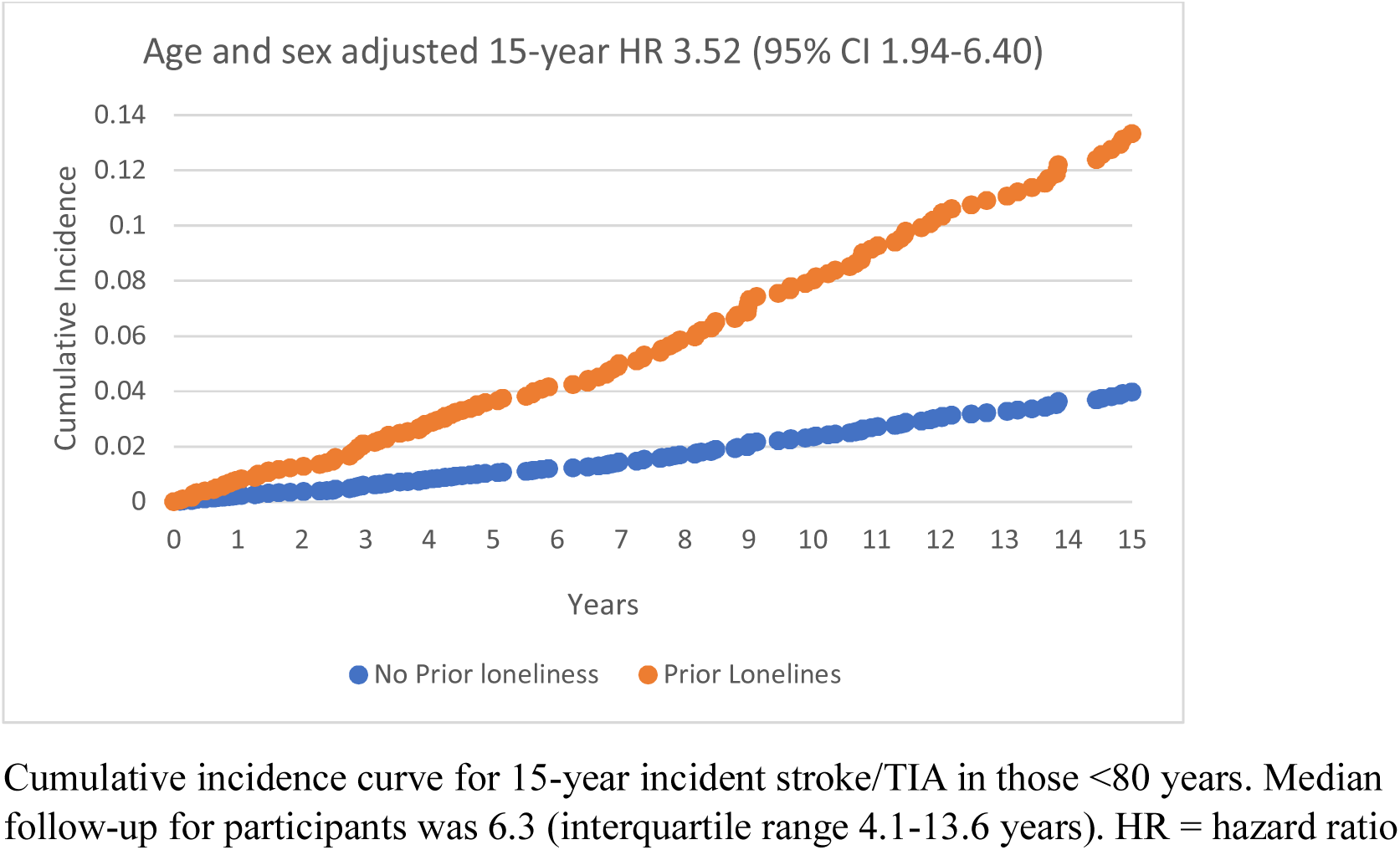
Fifteen-year Cumulative Incidence of stroke/TIA in those with and without prior loneliness defined as ≥3days/week, among participants aged <80 years

**Supplementary Table 1A:** Distribution of participants with loneliness pattern and stroke.

| Distribution of loneliness Change and Stroke/TIA cases with loneliness defined as ≥ 3days/week |  |  |  |  |  |  |
|---|---|---|---|---|---|---|
|  | Stroke/TIA cases |  |  |  |  |  |
|  | All participants |  | <80 years |  | ≥80 years |  |
|  | Stroke/TIA cases | Total number | Stroke/TIA cases | Total number | Stroke/TIA cases | Total number |
| No loneliness | 177 | 3944 | 119 | 3622 | 58 | 322 |
| Incident | 7 | 186 | 6 | 160 | 1 | 26 |
| Transient | 10 | 137 | 8 | 119 | 2 | 18 |
| Persistent | 4 | 40 | 4 | 35 | 0 | 5 |
| Distribution of loneliness and Stroke/TIA cases with loneliness defined as ≥ 1day/week |  |  |  |  |  |  |
|  | All participants |  | <80 years |  | ≥80 years |  |
|  | Stroke/TIA cases | Total number | Stroke/TIA cases | Total number | Stroke/TIA cases | Total number |
| No loneliness | 131 | 3104 | 90 | 2860 | 41 | 244 |
| Incident | 28 | 499 | 18 | 435 | 10 | 64 |
| Transient | 20 | 398 | 15 | 366 | 5 | 32 |
| Persistent | 19 | 306 | 14 | 275 | 5 | 31 |

**Supplementary Table 1B:** Distribution of participants with Prior and Baseline loneliness and stroke.

| Distribution of loneliness (Prior and Baseline) and Stroke/TIA cases with loneliness defined as ≥ 3days/week |  |  |  |  |  |  |
|  | Stroke/TIA cases |  |  |  |  |  |
|  | All participants |  | <80 years |  | ≥80 years |  |
|  | Stroke/TIA cases | Total number | Stroke/TIA cases | Total number | Stroke/TIA cases | Total number |
|---|---|---|---|---|---|---|
| Prior loneliness | 14 | 177 | 12 | 154 | 2 | 23 |
| No prior loneliness | 184 | 4130 | 125 | 3782 | 59 | 348 |
| Baseline loneliness | 11 | 226 | 10 | 195 | 1 | 31 |
| No baseline loneliness | 187 | 4081 | 127 | 3741 | 60 | 340 |
| Distribution of loneliness and Stroke/TIA cases with loneliness defined as ≥ 1day/week |  |  |  |  |  |  |
|  | All participants |  | <80 years |  | ≥80 years |  |
|  | Stroke/TIA cases | Total number | Stroke/TIA cases | Total number | Stroke/TIA cases | Total number |
| Prior loneliness | 39 | 704 | 29 | 641 | 10 | 63 |
| No prior loneliness | 159 | 3603 | 108 | 3295 | 51 | 308 |
| Baseline loneliness | 47 | 805 | 32 | 710 | 15 | 95 |
| No baseline loneliness | 151 | 3502 | 105 | 3226 | 46 | 226 |

**Supplementary Table 2:**
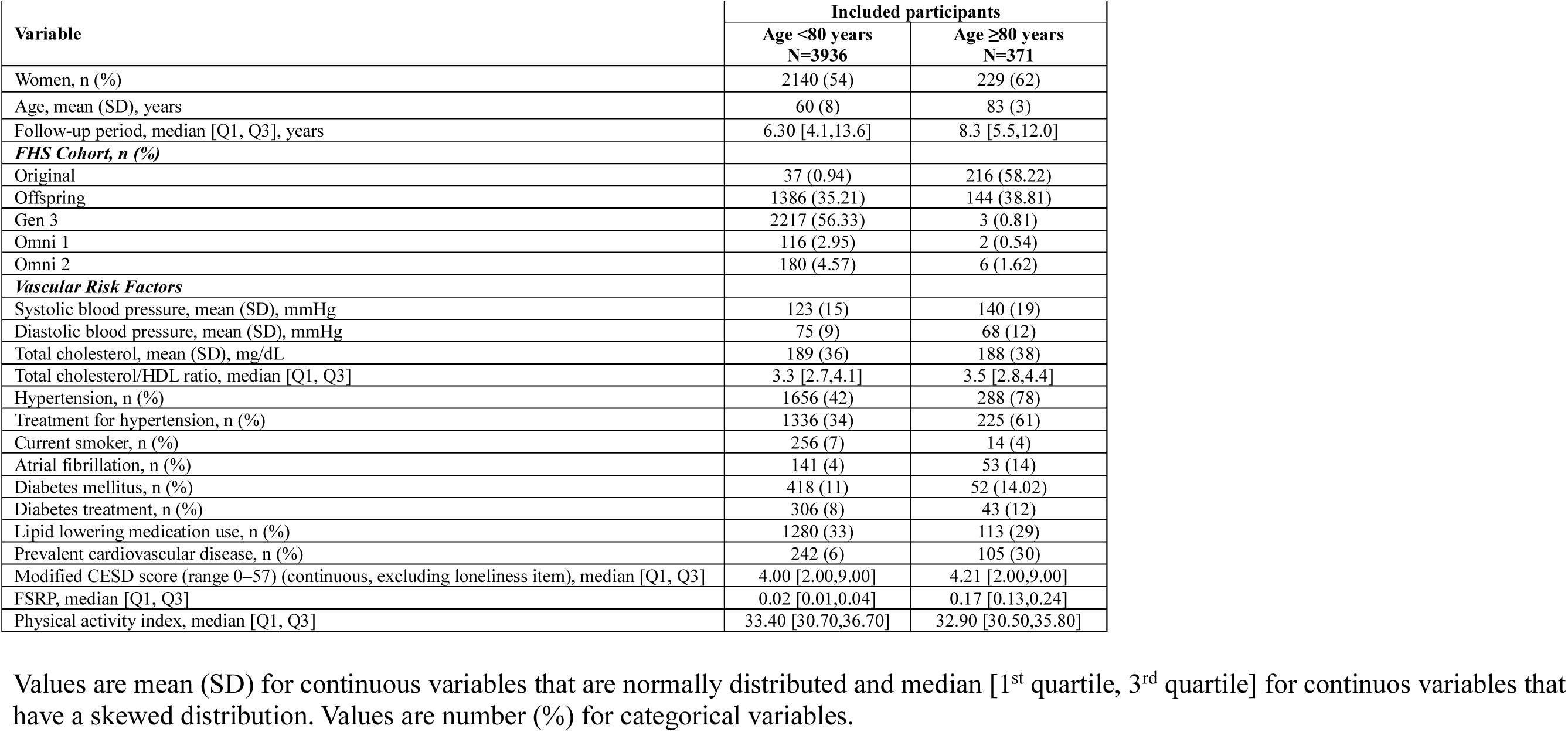
Baseline descriptives of Study participants categorized by age <80 and ≥80 years.

**Supplementary table 3:** Cox-proportional hazard regression of incident stroke by loneliness status ≥3 days/week (age ≥80 years).

|  | **Model 1 (n=371)** |  |  |
|---|---|---|---|
|  | ***Stroke/TIA cases/n*** | ***HR (95% CI)*** | ***P-value*** |
| Prior Loneliness |  |  |  |
| No | 59/348 | 1.00 (ref) |  |
| Yes | 2/23 | 0.60 (0.14,2.47) | ns |
| Baseline Loneliness |  |  |  |
| No | 60/340 | 1.00 (ref) |  |
| Yes | 1/31 | 0.21 (0.03,1.51) | ns |

Model 1: adjusted for age and sex

**Supplementary table 4:** Cox-proportional hazard regression of incident stroke by loneliness status ≥1 day/week (all participants).

|  | **Model 1 (n=4307)** |  |  | **Model 2 (n=4307)** |  |  | **Model 3 (n=4307)** |  |  | **Model 4 (n=4009)<sup>#</sup>** |  |  |
|---|---|---|---|---|---|---|---|---|---|---|---|---|
|  | ***Stroke/TIA cases/n*** | ***HR (95% CI)*** | ***P-value*** | ***Stroke/TIA cases/n*** | ***HR (95% CI)*** | ***P-value*** | ***Stroke/TIA cases/n*** | ***HR (95% CI)*** | ***P-value*** | ***Stroke/TIA cases/n*** | ***HR (95% CI)*** | ***P-value*** |
| Prior Loneliness |  |  |  |  |  |  |  |  |  |  |  |  |
| No | 159/3603 | 1.00 (ref) |  | 159/3603 | 1.00 (ref) |  | 159/3603 | 1.00 (ref) |  | 121/3355 | 1.00 (ref) |  |
| Yes | **39/704** | **1.51 (1.06,2.15)** | **0.022** | **39/704** | **1.51 (1.06,2.15)** | **0.022** | **39/704** | **1.84 (1.05,3.23)** | **0.033** | **32/654** | **1.60 (1.06,2.41)** | **0.024** |
| Baseline Loneliness |  |  |  |  |  |  |  |  |  |  |  |  |
| No | 151/3502 | 1.00 (ref) |  | 151/3502 | 1.00 (ref) |  | 151/3502 | 1.00 (ref) |  | 118/3281 | 1.00 (ref) |  |
| Yes | 47/805 | 1.18 (0.85,1.64) | 0.320 | 47/805 | 1.16 (0.84,1.62) | 0.368 | 47/805 | 0.78 (0.42,1.47) | 0.444 | 35/728 | 1.03 (0.67,1.59) | 0.896 |

Model 1: adjusted for age and sex
Model 2: adjusted for age, sex, Framingham Stroke Risk Profile Score (FSRP)
Model 3: adjusted for age, sex, FSRP, modified Center for Epidemiologic Studies Depression Scale (continuous, logarithm of CES-D excluding loneliness item)
Model 4: adjusted for adjusted for age, sex, FSRP, modified CES-D, physical activity index)
#model 4 done in offspring, third-generation, Omni 1 and 2 cohorts

**Supplementary table 5:** Cox-proportional hazard regression of incident stroke by loneliness status ≥1 day/week (age < 80 years).

|  | **Model 1 (n=3936)** |  |  | **Model 2 (n=3936)** |  |  | **Model 3 (n=3936)** |  |  | **Model 4 (n=3865)<sup>#</sup>** |  |  |
|---|---|---|---|---|---|---|---|---|---|---|---|---|
|  | ***Stroke/TIA cases/n*** | ***HR (95% CI)*** | ***P-value*** | ***Stroke/TIA cases/n*** | ***HR (95% CI)*** | ***P-value*** | ***Stroke/TIA cases/n*** | ***HR (95% CI)*** | ***P-value*** | ***Stroke/TIA cases/n*** | ***HR (95% CI)*** | ***P-value*** |
| Prior Loneliness |  |  |  |  |  |  |  |  |  |  |  |  |
| No | 108/3295 | 1.00 (ref) |  | 108/3295 | 1.00 (ref) |  | 108/3295 | 1.00 (ref) |  | 97/3232 | 1.00 (ref) |  |
| Yes | **29/641** | **1.76 (1.17,2.67)** | **0.007** | **29/641** | **1.76 (1.17,2.66)** | **0.007** | **29/641** | **3.17 (1.70,5.89)** | **<0.001** | **28/633** | **1.69 (1.08,2.64)** | **0.020** |
| Baseline Loneliness |  |  |  |  |  |  |  |  |  |  |  |  |
| No | 105/3226 | 1.00 (ref) |  | 105/3226 | 1.00 (ref) |  | 105/3226 | 1.00 (ref) |  | 96/3173 | 1.00 (ref) |  |
| Yes | 32/710 | 1.26 (0.85,1.88) | 0.253 | 32/710 | 1.25 (0.84,1.85) | 0.279 | 32/710 | 1.17 (0.59,2.31) | 0.652 | 29/692 | 1.06 (0.66,1.70) | 0.818 |

Model 1: adjusted for age and sex
Model 2: adjusted for age, sex, Framingham Stroke Risk Profile score (FSRP)
Model 3: adjusted for age, sex, FSRP, modified Center for Epidemiologic Studies Depression Scale (continuous, logarithm of CES-D excluding loneliness item)
Model 4: adjusted for adjusted for age, sex, FSRP, modified CES-D, physical activity index
#model 4 done in Offspring, Third Generation, Omni 1 and 2 cohorts

**Supplementary table 6:** Cox-proportional hazard regression of incident stroke by loneliness status ≥1 day/week (age ≥ 80 years).

|  | **Model 1 (n=371)** |  |  | **Model 2 (n=371)** |  |  | **Model 3 (n=371)** |  |  | **Model 4 (n=144)<sup>#</sup>** |  |  |
|---|---|---|---|---|---|---|---|---|---|---|---|---|
|  | ***Stroke/TIA cases/n*** | ***HR (95% CI)*** | ***P-value*** | ***Stroke/TIA cases/n*** | ***HR (95% CI)*** | ***P-value*** | ***Stroke/TIA cases/n*** | ***HR (95% CI)*** | ***P-value*** | ***Stroke/TIA cases/n*** | ***HR (95% CI)*** | ***P-value*** |
| Prior Loneliness |  |  |  |  |  |  |  |  |  |  |  |  |
| No | 51/308 | 1.00 (ref) |  | 51/308 | 1.00 (ref) |  | 51/308 | 1.00 (ref) |  | 24/123 | 1.00 (ref) |  |
| Yes | 10/63 | 1.14 (0.57,2.27) | ns | 10/63 | 1.14 (0.57,2.28) | ns | 10/63 | 0.60 (0.14,2.50) | ns | 4/21 | 1.08 (0.36,3.19) | ns |
| Baseline Loneliness |  |  |  |  |  |  |  |  |  |  |  |  |
| No | 46/276 | 1.00 (ref) |  | 46/276 | 1.00 (ref) |  | 46/276 | 1.00 (ref) |  | 22/108 | 1.00 (ref) |  |
| Yes | 15/95 | 1.16 (0.64,2.10) | ns | 15/95 | 1.10 (0.61,2.00) | ns | 15/95 | 0.20 (0.03,1.45) | ns | 6/36 | 0.94 (0.32,2.73) | ns |

Model 1: adjusted for age and sex
Model 2: adjusted for age, sex, Framingham Stroke Risk Profile score (FSRP)
Model 3: adjusted for age, sex, FSRP, modified Center for Epidemiologic Studies Depression Scale (continuous, logarithm of CES-D excluding loneliness item)
Model 4: adjusted for adjusted for age, sex, FSRP, modified CES-D, physical activity index
#model 4 done in Offspring, Third Generation, Omni 1 and 2 cohorts

